# Establishing Internationally Accepted Conceptual and Operational Definitions of Social Prescribing Through Expert Consensus: A Delphi Study

**DOI:** 10.1101/2022.11.14.22282098

**Authors:** Caitlin Muhl, Kate Mulligan, Imaan Bayoumi, Rachelle Ashcroft, Christina Godfrey

## Abstract

**Introduction:** With the social prescribing movement gaining traction globally, there is a need for an agreed definition of social prescribing. There are two types of definitions – conceptual and operational, meaning agreement on both types of definitions is needed.

**Objective:** The aim of this study was to establish internationally accepted conceptual and operational definitions of social prescribing.

**Design:** A three-round Delphi study was conducted.

**Methods:** Consensus was defined *a priori* as ≥80% agreement. In Round 1, participants were asked to list key elements that are essential to the conceptual definition of social prescribing and to provide corresponding statements that operationalize each of the key elements. In Round 2, participants were asked to rate their agreement with items from the first round for inclusion in the conceptual and/or operational definitions of social prescribing. Based on the findings from this round, the conceptual and operational definitions of social prescribing were developed, including long and short versions of the conceptual definition. In Round 3, participants were asked to rate their agreement with the conceptual and operational definitions of social prescribing.

**Participants:** This study involved an international, multidisciplinary panel of experts. The expert panel (n=48) represented 26 different countries across five continents, numerous expert groups, and a variety of years of experience with social prescribing, with the average being 5 years (range = 1-20 years).

**Results:** After three rounds, internationally accepted conceptual and operational definitions of social prescribing were established. The definitions were transformed into the Common Understanding of Social Prescribing (CUSP) conceptual framework.

**Conclusion:** This foundational work offers a common thread – a shared sense of what social prescribing is, which may be woven into social prescribing research, policy, and practice to foster common understanding of this concept.

**STRENGTHS AND LIMITATIONS OF THIS STUDY:**

- Strengths of this study include the consensus method that was chosen, the diversity and size of the expert panel, and the fact that consensus was defined *a priori* as ≥80% agreement
- Limitations of this study include the fact that only those who could speak, read, and write English were eligible to participate in this study, the attrition of the expert panel, and the fact that the expert panel did not reach 100% agreement on the definitions

## INTRODUCTION

Across the globe, the social prescribing movement is gaining momentum.[1] Morse et al[1] recently reported on global developments in social prescribing, which confirmed the existence of social prescribing initiatives in 17 countries, including Australia, Canada, China, Denmark, Finland, Germany, Ireland, Japan, the Netherlands, New Zealand, Portugal, Singapore, South Korea, Spain, Sweden, the United Kingdom (UK) (England, Scotland, Wales, and Northern Ireland), and the United States (US). Since then, the social prescribing movement has expanded its reach to include over 20 countries,[2] including Austria, Bosnia and Herzegovina, Brazil, Czech Republic, Ecuador, and Taiwan. The collective experience of these countries shows that social prescribing has the potential to support the achievement of global goals for health and wellbeing.[1]

While the term originates from the UK and dates back to almost a century ago,[1,3] the rise of social prescribing as a global phenomenon has mostly transpired within the past decade.[1] Throughout this period, there have been significant developments in the social prescribing movement. Across the globe, social prescribing organizations and networks have been established, including the Social Prescribing Network in 2015,[4] the Social Prescribing Youth Network in 2018,[5] the National Academy for Social Prescribing in 2019,[6] the World Health Organization and United Nations-linked Global Social Prescribing Alliance in 2021,[7] and the Canadian Institute for Social Prescribing in 2022.[8] Furthermore, annual events are held by the social prescribing community to celebrate social prescribing around the world, including the Social Prescribing Network Conference and Social Prescribing Day, which began in 2016 and 2019 respectively.[9,10] Additionally, a global network of student champions has emerged to build the social prescribing student movement – this began with the launch of the UK student group in 2017 and recently expanded to include student groups in Australia, Canada, Japan, Portugal, Singapore, and the US, which led to the creation of the Social Prescribing International Student Movement Framework in 2021[3] and the launch of the Global Social Prescribing Student Council in 2022. Most recently, as a way to foster the implementation of social prescribing, the World Health Organization released a social prescribing toolkit and online training module.[11,12]

Despite these developments in the social prescribing movement, an agreed definition of social prescribing has yet to emerge.[1,9,13–34] The reality is that the definition of social prescribing varies across and within countries.[1] Experts have deemed the concept to be nebulous and open to different interpretations.[13] The lack of consensus around the definition hinders efforts to generate robust evidence on social prescribing,[14,15] inhibits policy and practice development related to social prescribing,[15,17] and contributes to low public awareness of social prescribing.[25] Given how rapidly the social prescribing movement is growing and the different forms it is taking around the world,[1] there is a need for a common thread – a shared sense of what social prescribing is. This calls for an agreed definition of social prescribing.[14–17]

There are two types of definitions – conceptual and operational.[35] A conceptual definition outlines what a concept means but it does not explain how to measure it, whereas an operational definition outlines how to measure a concept but it does not explain what it means. Since there is a lack of agreement on both the conceptualization and operationalization of social prescribing,[14,33,34] agreement on both types of definitions is needed. The aim of this study was to establish internationally accepted conceptual and operational definitions of social prescribing.

## METHODS

### Study Design

A three-round Delphi study was conducted to establish internationally accepted conceptual and operational definitions of social prescribing with an international, multidisciplinary panel of experts. The Delphi technique is a method of gaining consensus on a particular topic through multiple rounds of questioning of experts in the field, who remain anonymous and receive feedback between each round.[36,37] This consensus method is widely used by health science researchers to achieve expert consensus,[38] particularly to establish agreed definitions.[39–75]

### Study Overview

This study was conducted between April 2022 and September 2022. Given the importance of the quality of the online survey platform to the success of this study, we carefully reviewed and tested several different options prior to selection. We used Welphi (www.welphi.com), which is an online survey platform that is specifically designed for Delphi studies. The Welphi team made coding changes when necessary to ensure that the online survey platform met the needs of this study.

Before the start of each round, we completed survey development and pilot testing. This consisted of building the survey on the online survey platform and subsequently completing a test version to make improvements. Participants received a notification email one week prior to the launch of each round. At the start of each round, participants received an email with a link to the survey and a link to a calendar with the details of the round. Participants had two weeks to complete each round. Each survey began with a welcome page, which provided an overview of the survey, described the data analysis procedures, explained what kind of feedback would be provided at the start of the next round, highlighted the aim of the study, and outlined important information about conceptual and operational definitions to ensure that there was common understanding of these terms. Participants were able to return to the survey as many times as they liked until the closure of the round – their progress was saved if they left the survey and came back at a later time, and even after completing the survey, they were able to make changes to their submission until the round closed. Each survey took approximately 30-60 minutes to complete. Reminder emails were sent out one week, two days, and one day before the closure of each round, as well as the day of the closure of each round. After two weeks, an email was sent out to non-responders to give them an additional three days to complete the survey – if they did not respond by that time, they were removed from the study. In other words, only those who completed Round 1 were eligible to participate in Round 2, and only those who completed Round 2 were eligible to participate in Round 3. After each round closed, we completed data analysis. This took place over three weeks in Round 1, two weeks in Round 2, and one day in Round 3.

### Participants

There is a lack of standard guidelines and agreement in the literature as to what constitutes an expert for Delphi studies.[36,37] For this study, experts were defined according to the following criteria: (1) Person involved with the Social Prescribing Network; or (2) Person involved with the Social Prescribing Youth Network; or (3) Person involved with the Global Social Prescribing Alliance; or (4) Person involved with the National Academy for Social Prescribing; or (5) Person involved with the Canadian Institute for Social Prescribing; or (6) Student involved with any national social prescribing student group; or (7) Author of academic or grey literature on social prescribing, even if not labelled as “social prescribing”; or (8) Researcher involved in social prescribing, even if not labelled as “social prescribing”; or (9) Health care provider involved in social prescribing, even if not labelled as “social prescribing”; or (10) Link worker involved in social prescribing, even if not labelled as “link worker” or “social prescribing”; or (11) Patient involved in social prescribing, even if not labelled as “social prescribing”; or (12) Health care administrator or manager tasked with overseeing the use of social prescribing, even if not labelled as “social prescribing”. Furthermore, since this study was in English, only those who could speak, read, and write English were eligible to participate in this study. Determination of eligibility was at the discretion of the interested party.

Although there is a lack of standard guidelines and agreement in the literature as to the appropriate size of the expert panel for Delphi studies,[36–38] the average number of experts that health science researchers include in Delphi studies is 40,[38] and most Delphi studies that have been conducted by health science researchers to establish agreed definitions have had between 20-60 people on the expert panel.[41–61] We therefore attempted to recruit approximately 40 participants (range = 20-60 participants).

### Recruitment

Targeted recruitment began in April 2022 when the registration survey for the study opened for members of the Global Social Prescribing Alliance. Following this, the first author (CM) developed a comprehensive list with the names and contact information of over 400 experts from across the globe, determined the top experts from each country by comparing their expertise in social prescribing, and sent out invitations to these individuals via email. All of this information was obtained from public sources. Throughout the recruitment process, the acquisition of an international, multidisciplinary panel of experts was prioritized. We used a matrix to ensure that there was diversity amongst experts in terms of country, job title, expertise in social prescribing, and years of experience with social prescribing. The registration survey was closely monitored to focus recruitment efforts on experts from underrepresented groups – expert groups with significantly lower representation than others. Open recruitment began in May 2022 when we sent out invitations to experts from underrepresented groups via relevant communication channels (i.e., Social Prescribing Network newsletter, Social Interventions Research and Evaluation Network listserv, Canadian Social Prescribing Community of Practice listserv) and advertised invitations to these experts via relevant social media platforms (i.e., Twitter, LinkedIn). Consistent with snowball sampling, experts were asked to disseminate the call to relevant contacts.

Experts received an email with a link to the registration survey, which was administered through Qualtrics (www.qualtrics.com). The first page of the survey was the Letter of Information. Experts were informed that by proceeding to the next page, they were consenting to participate in the study. Consent was voluntary, informed, and ongoing. Sociodemographic data, including name, email address, country, job title, organization, expertise in social prescribing, and years of experience with social prescribing, were collected through the registration survey. Participants were notified that the first round of the study would begin once enough experts had registered.

### Consensus

While there is a lack of standard guidelines and agreement in the literature as to what constitutes consensus for Delphi studies,[36–38,76] most Delphi studies define consensus as a certain percentage of participants being in agreement.[36] For this study, consensus was defined *a priori* as ≥80% agreement, meaning ≥80% of participants had to rate their agreement as Agree (4) or Strongly Agree (5) on a 5-point Likert scale (Strongly Disagree (1), Disagree (2), Neutral (3), Agree (4), Strongly Agree (5)). This threshold was chosen as an appropriate cutoff given that most health science researchers that have conducted Delphi studies to establish agreed definitions have defined consensus as either ≥70%, ≥75%, or ≥80% agreement,[40–55,66–74] with ≥80% being the most stringent level of agreement.

### Rounds

This study consisted of three rounds. This number was not set in advance, although we anticipated that this study would consist of 3-5 rounds. Recognizing that Delphi studies can consist of any number of rounds,[37] this prediction was not only based on the format of each round in this study, but also on the fact that Delphi studies commonly involve 3-5 rounds,[37] and that most Delphi studies that have been conducted by health science researchers to establish agreed definitions have consisted of 3-5 rounds.[39–43,47–60,70–74] However, we did set a maximum number of rounds in advance – if consensus was not reached on the definitions by a fifth round, then a sixth and final round would take place in the form of a meeting with participants via teleconferencing software to achieve consensus through discussion.

### Data Collection

The Delphi surveys were conducted over four months. Participants were automatically assigned randomly generated participant codes by the online survey platform, meaning all responses were anonymized.

#### Round 1

The first round was conducted between June 2022 and July 2022. Participants completed an open-ended survey to gather information through open-ended questions. The first question asked participants to list key elements that are essential to the conceptual definition of social prescribing. The second question built on this by asking participants to provide corresponding statements that operationalize each of the key elements.

#### Round 2

The second round was conducted between July 2022 and August 2022. Participants rated items from the first round through a structured survey. Participants did not have to consider the type of definition when rating the items. This protocol amendment was put in place to make this process as straightforward as possible.[77] The items were simply presented to participants, and they were asked to rate their agreement with each item for inclusion in the conceptual and/or operational definitions of social prescribing. Ratings were given on a 5-point Likert scale (Strongly Disagree (1), Disagree (2), Neutral (3), Agree (4), Strongly Agree (5)). Participants were reminded that items would be accepted for inclusion in the conceptual and/or operational definitions of social prescribing if ≥80% of participants rated their agreement to include them as Agree (4) or Strongly Agree (5). A free-text box was also provided to add comments about the items and/or participants’ rating of the items, and participants were encouraged to do so.

#### Round 3

The third round was conducted between August 2022 and September 2022. Participants completed a structured survey to rate their agreement with the conceptual and operational definitions of social prescribing, including long and short versions of the conceptual definition. Consistent with the second round, ratings were given on a 5-point Likert scale (Strongly Disagree (1), Disagree (2), Neutral (3), Agree (4), Strongly Agree (5)). Participants were reminded that consensus would be reached on the definitions if ≥80% of participants rated their agreement with them as Agree (4) or Strongly Agree (5). A free-text box was also provided to add comments about the definitions, and participants were encouraged to do so, particularly if they rated their agreement as Strongly Disagree (1), Disagree (2), or Neutral (3). Participants were informed about next steps – if consensus was not reached on all three definitions in this round, then the comments would be used to make modifications to any definition that did not reach consensus and another round would subsequently be completed, but if consensus was reached, then no further changes would be made to the definitions and no additional rounds would be completed.

### Data Analysis

Data collected from the registration survey were analyzed with Microsoft Excel (www.microsoft.com). For the Delphi surveys, the online survey platform generated the pooled results for each round. Quantitative data were expressed in percentages as statistical group response. Qualitative data were presented for each survey item and analyzed through qualitative content analysis, which is “an approach to text analysis which combines strict rulebound interpretive category assignments with quantifications of category occurrences” (p209).[41] This was done with QCAmap (www.qcamap.org), which is an online software program that is specifically designed for qualitative content analysis. QCAmap interactively guides users through the steps of qualitative content analysis,[78] thereby serving as a valuable tool for us to ensure that we successfully completed each step. After the first and second authors (CM and KM) coded the data, the results were compared to determine inter-coder agreement. Once the first author (CM) coded the data a second time, the results were compared to determine intra-coder agreement. The first author (CM) had to rate the agreement as bad, moderate, good, or excellent.

#### Round 1

Qualitative content analysis was conducted to analyze participants’ responses. Through inductive category formation, the first author (CM) organized the data into categories, which was subsequently reviewed by the second author (KM). There was excellent inter-coder (CM and KM) and intra-coder (CM) agreement. The findings were used to create a structured survey for the next round. At the start of the next round, participants received feedback from this round, which consisted of a summary of participants’ responses. All responses were anonymized.

#### Round 2

For the quantitative data, the first author (CM) examined the pooled results that were generated by the online survey platform. Items where ≥80% of participants rated their agreement to include them as Agree (4) or Strongly Agree (5) were accepted for inclusion in the conceptual and/or operational definitions of social prescribing. Items where the percentage of agreement was within 2% of the 80% threshold, meaning 78% of participants rated their agreement to include them as Agree (4) or Strongly Agree (5), were also accepted for inclusion in the conceptual and/or operational definitions of social prescribing. This protocol amendment was put in place to err on the side of caution, which reflects the pragmatic nature of this work.[77] The remaining items were not accepted for inclusion in the conceptual or operational definitions of social prescribing, as ≤75% of participants rated their agreement to include them as Agree (4) or Strongly Agree (5). As for the qualitative data, qualitative content analysis was conducted to analyze participants’ responses. Through inductive category formation, the first author (CM) organized the data into categories, which was subsequently reviewed by the second author (KM). There was excellent inter-coder (CM and KM) and intra-coder (CM) agreement. Based on the findings from this round, the conceptual and operational definitions of social prescribing were developed, including long and short versions of the conceptual definition. At the start of the next round, participants received feedback from this round. Quantitative feedback consisted of the percentage of agreement and the individual response of each participant in relation to the group response, and qualitative feedback consisted of a summary of participants’ responses. All responses were anonymized.

#### Round 3

For the quantitative data, the first author (CM) examined the pooled results that were generated by the online survey platform, which revealed that ≥80% of participants rated their agreement as Agree (4) or Strongly Agree (5) for all three definitions. As for the qualitative data, it was not necessary to conduct qualitative content analysis due to the quantitative findings.

### Quality and Transparency

This study has been conducted and reported in accordance with the Conducting and REporting DElphi Studies (CREDES) guideline.[79] A protocol has been registered on Open Science Framework (osf.io/pfyqg) and submitted for publication[77] to promote transparency.

### Patient and Public Involvement

There was patient and public involvement on the expert panel. However, there was no patient or public involvement in the design, conduct, reporting, or dissemination plans of this research.

### Ethics

This study has been reviewed for ethical compliance by the Queen’s University Health Sciences and Affiliated Teaching Hospitals Research Ethics Board (NURS-540-22).

## RESULTS

### Participants

Altogether, 58 experts registered to participate in this study. As depicted in Figure 1, 48/58 experts (83%) completed Round 1, 40/48 experts (83%) completed Round 2, and 37/40 experts (93%) completed Round 3. The sociodemographic characteristics of the expert panel, meaning those who participated in one or more rounds of the study, are presented in Table 1. The expert panel represented 26 different countries across five continents, which is illustrated in Figure 2. A significant proportion of participants (40%) represented countries in the UK (n=14) and Canada (n=5), which are considered leaders in social prescribing and are the only places in the world with national social prescribing organizations. However, the majority of participants (60%) represented other countries (n=29), which reflects the growth of the social prescribing movement and demonstrates our efforts to recruit an international panel of experts. Beyond the diversity of nationalities amongst members of the expert panel, there was also diversity in terms of expertise in social prescribing, with representation from every type of expert group solicited. Researchers involved in social prescribing (n=29) and authors of academic or grey literature on social prescribing (n=25) had the greatest representation. There was also diversity in terms of years of experience with social prescribing, with the average being 5 years (range = 1-20 years).

**Table 1.** Sociodemographic characteristics of the expert panel.

| Sociodemographic Characteristic | Expert Panel (n=48) |
| --- | --- |
| **Country** |  |
| *Asia* |  |
| China | 1 (2%) |
| Japan | 1 (2%) |
| Singapore | 2 (4%) |
| South Korea | 1 (2%) |
| Taiwan | 1 (2%) |
| *Europe* |  |
| Austria | 1 (2%) |
| Bosnia and Herzegovina | 2 (4%) |
| Czech Republic | 1 (2%) |
| Denmark | 1 (2%) |
| England | 11 (23%) |
| Finland | 2 (4%) |
| Germany | 1 (2%) |
| Ireland | 1 (2%) |
| Netherlands | 1 (2%) |
| Northern Ireland | 1 (2%) |
| Portugal | 2 (4%) |
| Scotland | 1 (2%) |
| Spain | 1 (2%) |
| Sweden | 2 (4%) |
| Wales | 1 (2%) |
| *North America* |  |
| Canada | 5 (10%) |
| United States | 2 (4%) |
| *Oceania* |  |
| Australia | 3 (6%) |
| New Zealand | 1 (2%) |
| *South America* |  |
| Brazil | 1 (2%) |
| Ecuador | 1 (2%) |
| **Expertise in Social Prescribing** |  |
| Person involved with the Social Prescribing Network | 17 (35%) |
| Person involved with the Social Prescribing Youth Network | 4 (8%) |
| Person involved with the Global Social Prescribing Alliance | 5 (10%) |
| Person involved with the National Academy for Social Prescribing | 7 (15%) |
| Person involved with the Canadian Institute for Social Prescribing | 2 (4%) |
| Student involved with a national social prescribing student group | 4 (8%) |
| Author of academic or grey literature on social prescribing* | 25 (52%) |
| Researcher involved in social prescribing* | 29 (60%) |
| Health care provider involved in social prescribing* | 12 (25%) |
| Link worker involved in social prescribing*† | 8 (17%) |
| Patient involved in social prescribing* | 1 (2%) |
| Health care administrator or manager tasked with overseeing the use of social prescribing* | 8 (17%) |
| **Years of Experience with Social Prescribing** |  |
| 1-3 | 18 (38%) |
| 4-6 | 15 (31%) |
| 7-9 | 8 (17%) |
| 10+ | 7 (15%) |

\*Includes those who do not label it as “social prescribing”
†Includes those who do not label it as “link worker”

**Figure 1.**
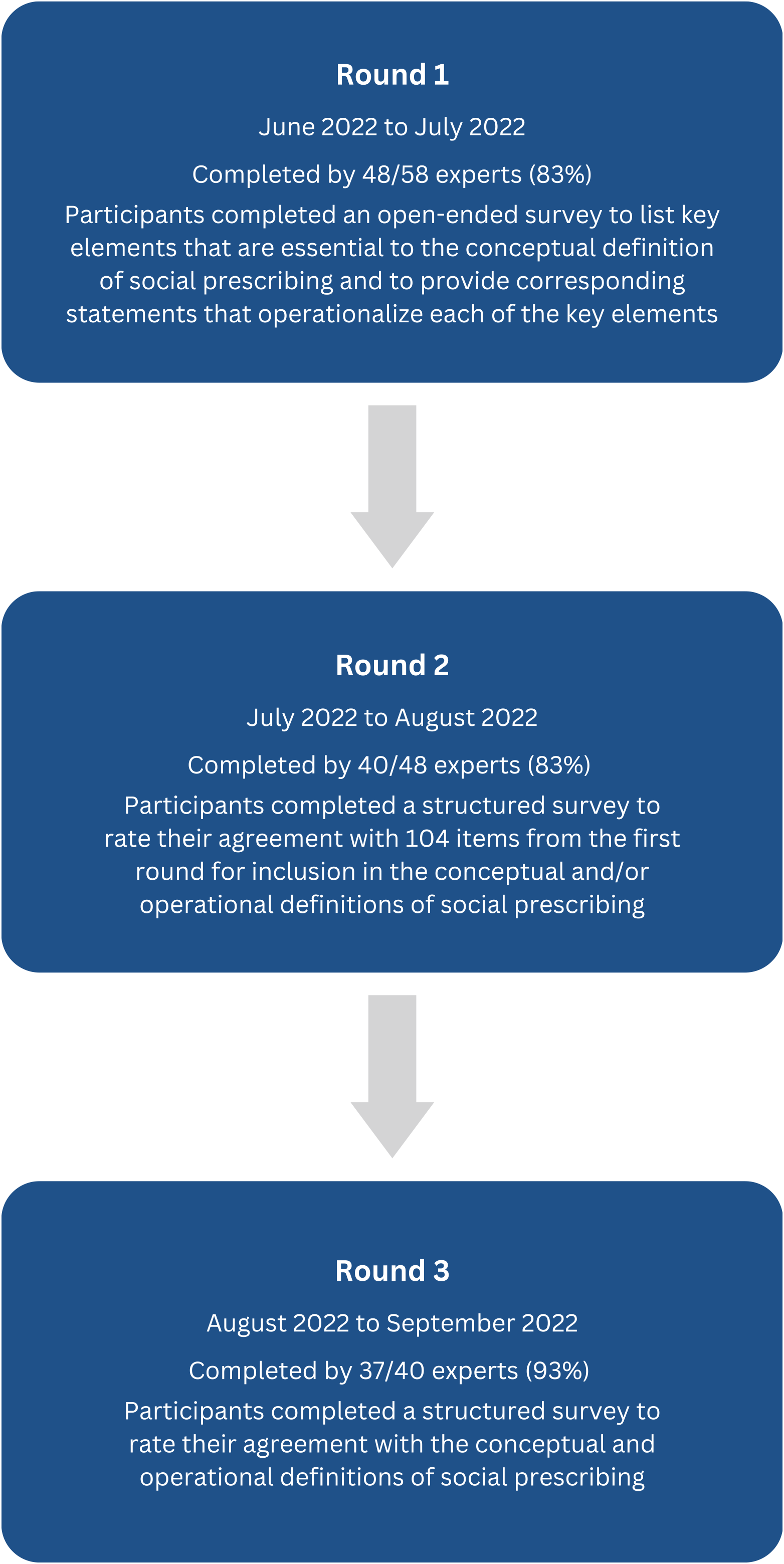
Delphi study flow chart.

**Figure 2.**
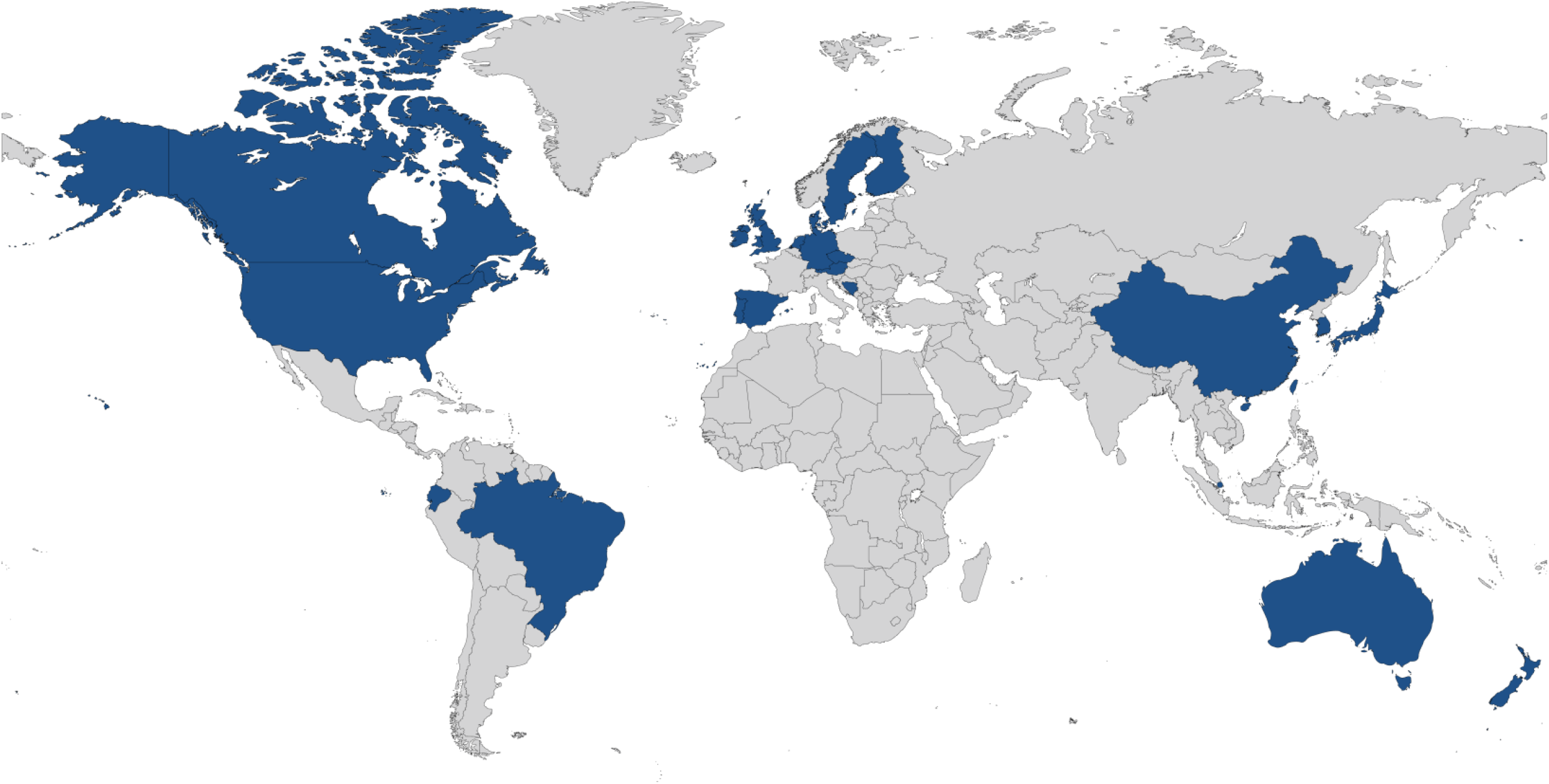
Countries represented by the expert panel: (1) Asia: China, Japan, Singapore, South Korea, and Taiwan; (2) Europe: Austria, Bosnia and Herzegovina, Czech Republic, Denmark, England, Finland, Germany, Ireland, the Netherlands, Northern Ireland, Portugal, Scotland, Spain, Sweden, Wales; (3) North America: Canada and the United States; (4) Oceania: Australia and New Zealand; and (5) South America: Brazil and Ecuador.

### Round 1

When asked to list key elements that are essential to the conceptual definition of social prescribing, the expert panel submitted 207 responses, which ranged in length from 1-112 words. When asked to provide corresponding statements that operationalize each of the key elements, the expert panel submitted 179 responses, which ranged in length from 1-411 words. Qualitative content analysis resulted in a list of 305 items. Outliers were removed from the list, meaning only those items that were mentioned by >1 participant were retained to bring forward to the second round, which resulted in a final list of 104 items. These items were divided into four groups: (1) Purpose (6 items); (2) People (29 items); (3) Properties (30 items); and (4) Process (39 items). The final list of items from this round is presented in Online Supplemental Material 1.

### Round 2

The results from this round are presented in Online Supplemental Material 1. Out of 104 items, there were 63 items in which ≥80% of participants rated their agreement to include them as Agree (4) or Strongly Agree (5). These items were accepted for inclusion in the conceptual and/or operational definitions of social prescribing. For six items, the percentage of agreement was within 2% of the 80% threshold, meaning 78% of participants rated their agreement to include them as Agree (4) or Strongly Agree (5). These items were also accepted for inclusion in the conceptual and/or operational definitions of social prescribing. For the remaining 35 items, ≤75% of participants rated their agreement to include them as Agree (4) or Strongly Agree (5).

These items were not accepted for inclusion in the conceptual or operational definitions of social prescribing. As for the qualitative data, 134 comments were submitted about the items and participants’ rating of the items, with no comments for 47 items, 1 comment for 24 items, and >1 comment for 33 items. Qualitative content analysis resulted in a list of suggestions for the conceptual and operational definitions of social prescribing. Consistent with the first round, outliers were removed from the list, meaning only those suggestions that were made by >1 participant were retained for inclusion in the definitions. Furthermore, only those suggestions that were deemed relevant based on the quantitative findings from this round were retained for inclusion in the definitions. This resulted in a final list of five suggestions: (1) Social prescribing offers a way to mitigate the impacts of adverse social determinants of health and health inequities, but does not address them; (2) Social prescribing not only takes place in clinical settings but also in community settings; (3) The connector supports the person to access community resources, but does not always support them over the long term; (4) The term ‘personalized support’ should be used instead of the term ‘personalized care’; and (5) Monitoring and evaluation not only includes the impact on the person and clinical setting but also on non-clinical supports and services and the community. Based on the findings from this round, the conceptual and operational definitions of social prescribing were developed, including long and short versions of the conceptual definition. The definitions are presented in Table 2.

**Table 2.**
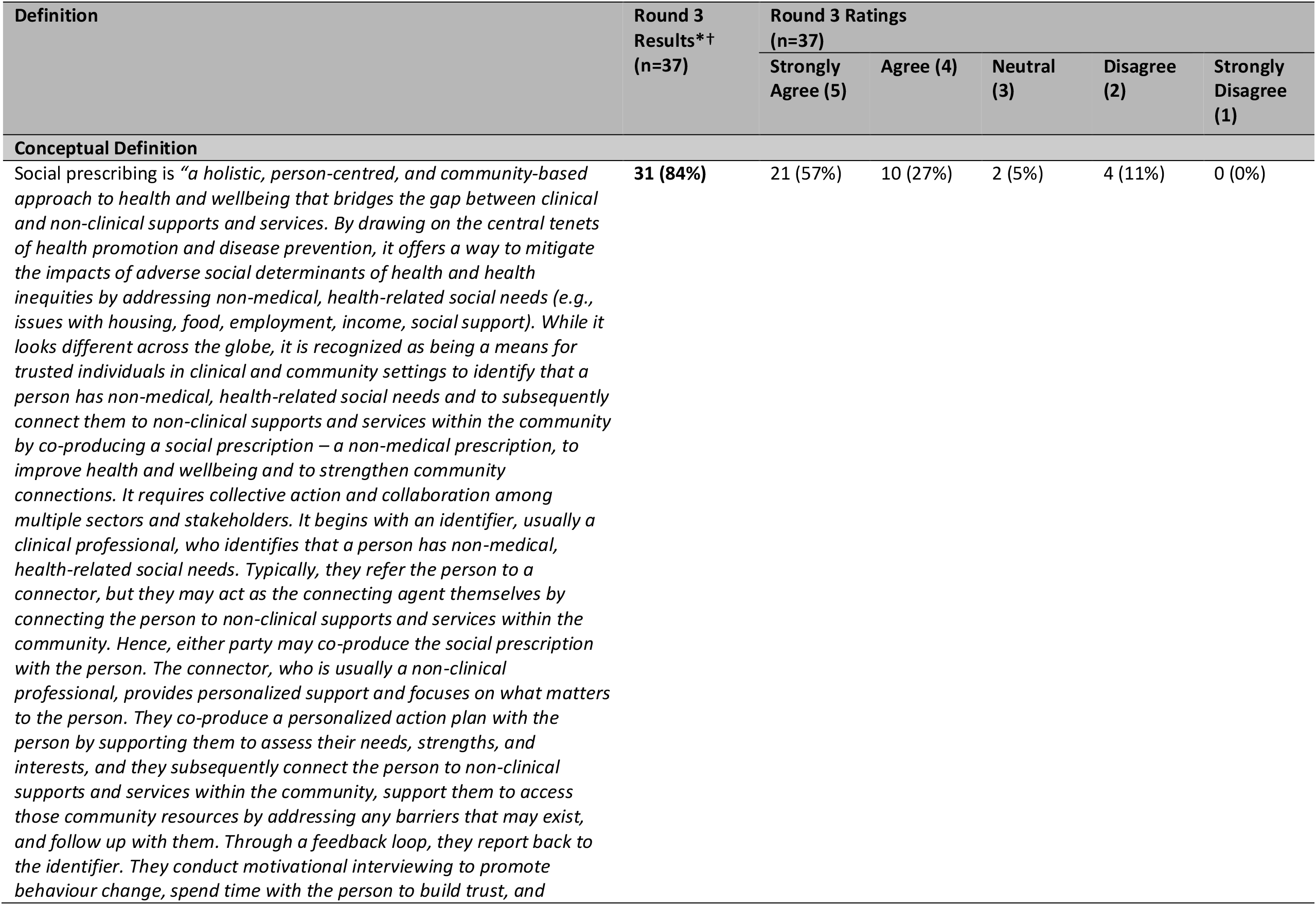
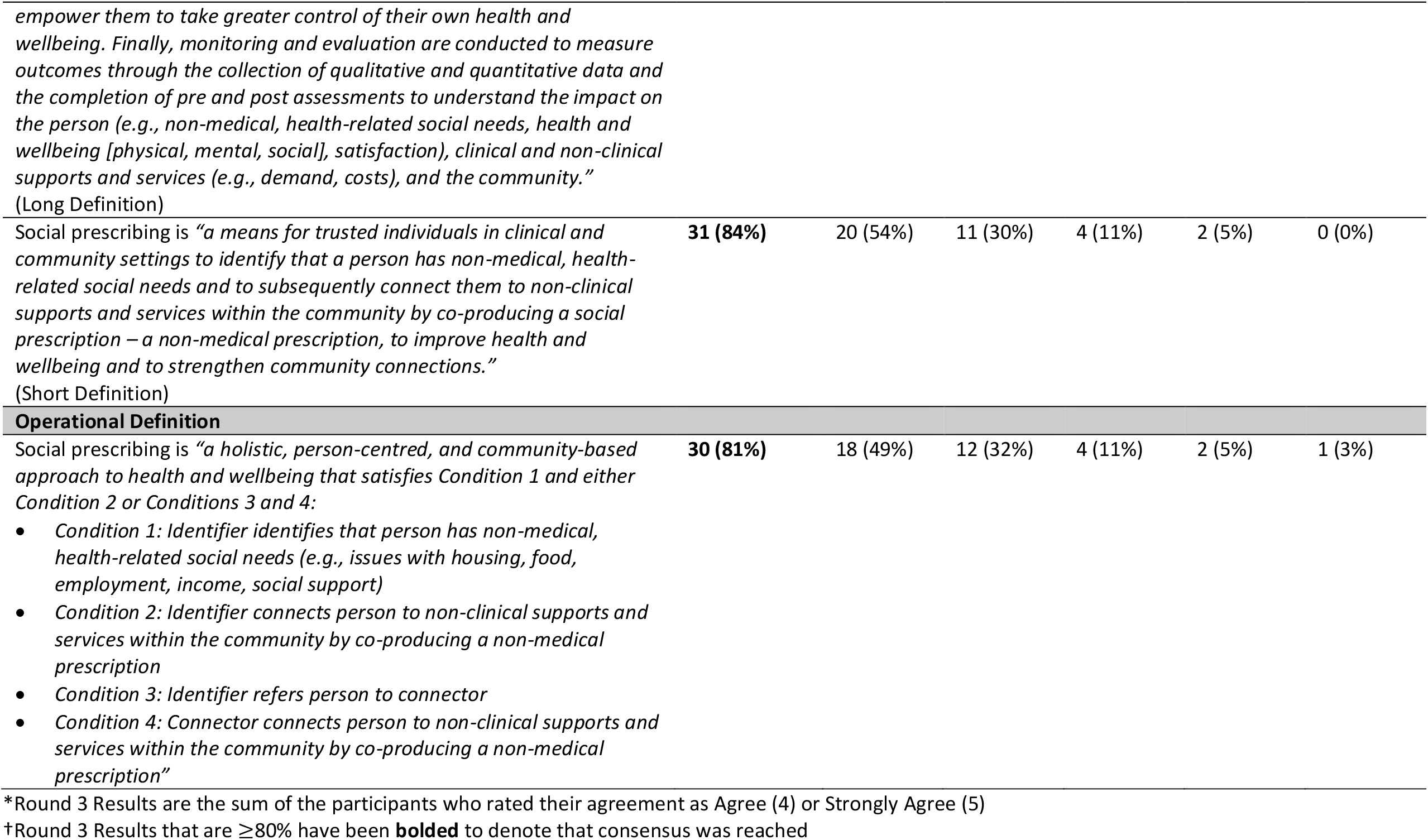
Conceptual and operational definitions of social prescribing.

### Round 3

The results from this round are presented in Table 2. Out of 37 participants, 84% (n=31) rated their agreement with the long version of the conceptual definition as Agree (4) or Strongly Agree (5), 84% (n=31) rated their agreement with the short version of the conceptual definition as Agree (4) or Strongly Agree (5), and 81% (n=30) rated their agreement with the operational definition as Agree (4) or Strongly Agree (5), meaning consensus was reached on all three definitions. As for the qualitative data, seven comments were submitted for each definition. However, it was not necessary to conduct qualitative content analysis due to the quantitative findings. Thus, no further changes were made to any of the definitions. We notified participants that consensus was reached on the definitions and that the study was complete, which signified the successful development of internationally accepted conceptual and operational definitions of social prescribing.

### Conceptual Framework

Upon completion of the study, we transformed the definitions into the Common Understanding of Social Prescribing (CUSP) conceptual framework, which is presented in Figure 3. The CUSP acronym reflects the potential of this foundational work to bring about a point of transition in the social prescribing movement through the advancement of common understanding.

**Figure 3.**
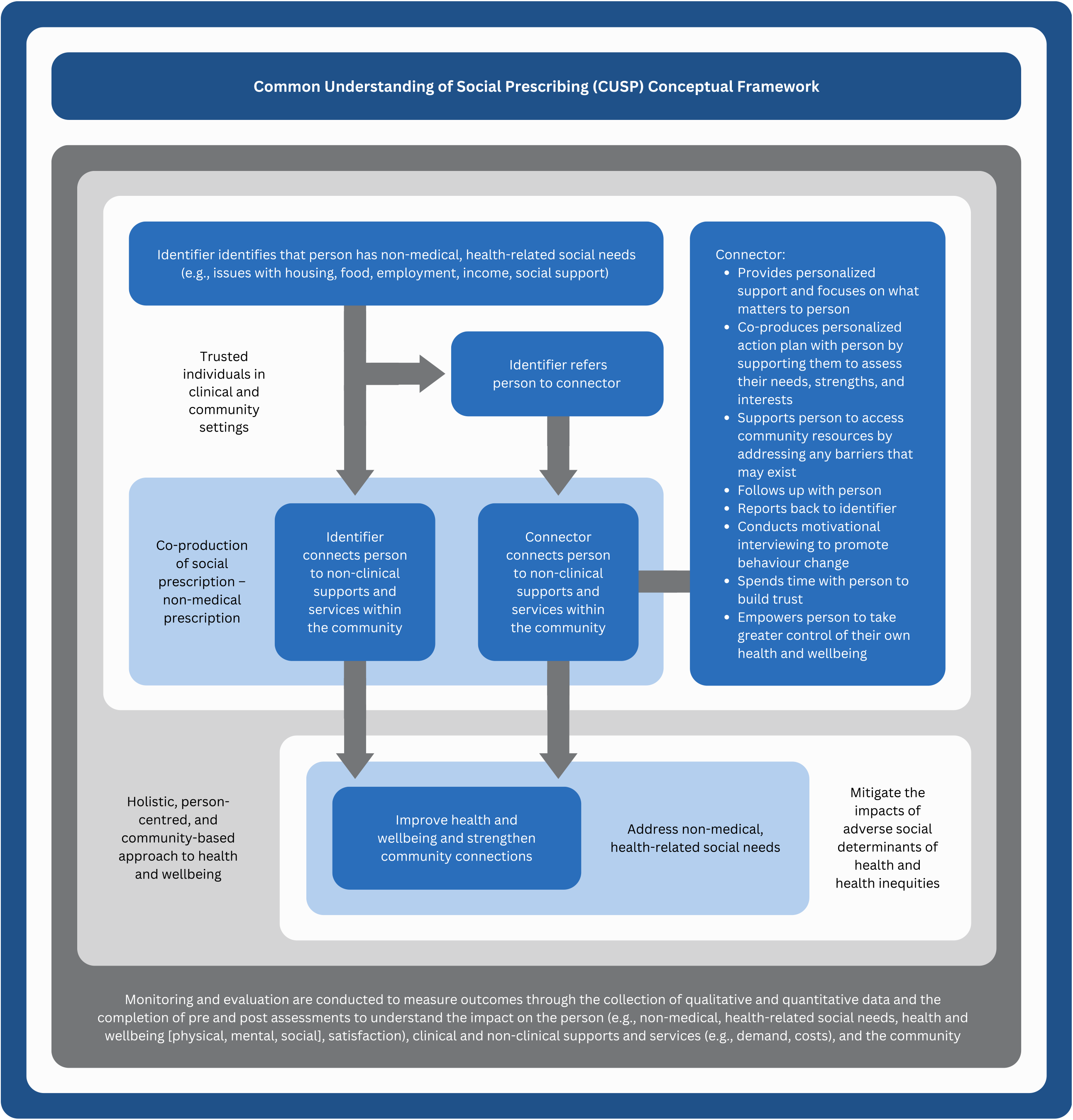
Common Understanding of Social Prescribing (CUSP) conceptual framework.

## DISCUSSION

For the first time in the history of the social prescribing movement, we now have internationally accepted conceptual and operational definitions of social prescribing. In addition, the conceptual definitions are distinct from pre-existing definitions, and to our knowledge, the operational definition is the first in the world. The definitions are flexible yet concrete – they recognize that social prescribing looks different around the world and account for those nuances, but at the same time, they outline the structural components of social prescribing that are shared across the globe. Along with the CUSP conceptual framework, the definitions provide a shared sense of what social prescribing is and offer a means of fostering common understanding of this concept.

The definitions and framework correspond to the evidence base on social prescribing and related concepts. Up to this point, it has been unclear as to who the social prescriber is[17] and the stage at which the social prescription occurs.[80] The outputs of this work help to clarify these core components of social prescribing, with the operational definition providing the clearest example of this – Condition 2 is the stage at which the social prescription occurs when the identifier is the social prescriber, whereas Condition 4 is the stage at which the social prescription occurs when the connector is the social prescriber. Additionally, the outputs of this work reflect current understanding of key facets of social prescribing, such as the holistic approach,[7,11] the central role that is played by identifying non-medical, health-related social needs and subsequently connecting to non-clinical supports and services within the community,[80,81] the linkages to improving health and wellbeing[1,7,11] and to strengthening community connections,[82] the responsibilities of the connector,[7,11,12,30,81] and the importance of monitoring and evaluation.[11,12,30] From a global health standpoint, Morse et al[1] highlight that social prescribing links to multiple trends in global health care, many of which are found in the outputs of this work, such as person-centredness, co-production, and health promotion. Furthermore, the well-known relationship between adverse social determinants of health, health inequities, and non-medical, health-related social needs is depicted in the outputs of this work.[83–86]

Meaningful insights are gleaned by comparing and contrasting the definitions with pre-existing definitions. Malby et al[13] state that the most common denominator among different definitions of social prescribing is the non-clinical aspect, which is a key element of the definitions established in this study. The most widely used definition of social prescribing comes from the Social Prescribing Network: “enabling health care professionals to refer patients to a link worker, to co-design a non-clinical social prescription to improve their health and wellbeing” (p19).[9] The latter half of this definition bears a resemblance to the following excerpt from the conceptual definitions established in this study: “…co-producing a social prescription – a non-medical prescription, to improve health and wellbeing…”. Beyond this, however, the definitions differ in several important ways. Unlike the definitions established in this study, the Social Prescribing Network definition[9] situates social prescribing within clinical settings, as it labels the person as a patient and indicates that social prescribing always begins with a health care professional. This is a common feature across pre-existing definitions of social prescribing, with one example being the definition that was recently developed by the World Health Organization: “a means for health care workers to connect patients to a range of non-clinical services in the community to improve health and wellbeing” (p2).[11] On the contrary, the expert panel in this study did not agree with the idea that social prescribing is limited to clinical settings, that the person is always a patient, or that social prescribing always begins with a health care professional – the percentage of agreement for the corresponding items was 38%, 43%, and 58% respectively (See Online Supplemental Material 1). As such, the definitions established in this study acknowledge that social prescribing not only takes place in clinical settings but also in community settings. Therefore, the person is not referred to as a patient, the term ‘care’ is not used, and the people involved are referred to as trusted individuals in clinical *and* community settings, which takes into account that the identifier is not always a clinical professional. Additionally, the definitions established in this study recognize the ways in which social prescribing relates to the social determinants of health, health equity, and non-medical, health-related social needs – this is noticeably absent from pre-existing definitions, including the Social Prescribing Network[9] and World Health Organization[11] definitions. Another important distinction is the universal language that was carefully chosen for the definitions established in this study to ensure that they would be applicable to all contexts. For example, the definitions refer to the person in the connector role as the connector, whereas this person is labelled as a ‘link worker’ in the Social Prescribing Network definition,[9] which limits the universality of this definition – Tierney et al[87] found that this is just one of 75 different terms that are used for the connector role in the UK alone. The definitions established in this study also include different variations of social prescribing and align with current understanding of this concept. The oft-cited Kimberlee[23] report outlines four different models of social prescribing: (1) Signposting; (2) Light; (3) Medium; and (4) Holistic. Based on recent interpretations of these models,[80,88] it is apparent that the latter three models are present in the definitions, with the operational definition providing the clearest example of this – Conditions 1 and 2 reflect the light model, whereas Conditions 1, 3, and 4 reflect the medium and holistic models. However, the signposting model is absent from the definitions, which reflects a shift in thinking by the social prescribing community and aligns with the argument recently brought forth by Morse et al[1] that signposting is distinct from social prescribing. Furthermore, the expert panel in this study did not agree with the idea that social prescribing always involves a connector – the percentage of agreement for the corresponding item was 45% (See Online Supplemental Material 1). Unlike the Social Prescribing Network definition,[9] which implies that a connector is always involved and therefore only accounts for the medium and holistic models, the definitions established in this study also account for the light model as they recognize that the identifier may act as the connecting agent themselves.

It is also necessary to consider the items that were not accepted for inclusion in the definitions and the implications of this for the social prescribing movement. Of particular significance are the items that are known to be contentious topics amongst members of the social prescribing community, such as the idea that social prescribing is an intervention or pathway, that social prescribing may be accessed through self-referral, and that one of the purposes of social prescribing is to reduce health care demand. The percentage of agreement for these items was 65%, 73%, and 55% respectively (See Online Supplemental Material 1). Although there was disagreement on these items, this did not preclude us from achieving the aim of this study.

There are several strengths and limitations of this study. One of the strengths is the consensus method that was chosen. The Delphi technique is known to reduce bias in the process of gaining consensus due to its unique characteristics, namely participant anonymity, multiple rounds of questioning, and provision of feedback between each round.[36,38,76] Another strength is the diversity and size of the expert panel. With respect to diversity, we were successful in acquiring an international, multidisciplinary panel of experts. Not only does the heterogeneous nature of the expert panel increase the validity of the findings,[36] but it also means that the definitions are relevant to different countries and stakeholders. With respect to size, there were 48 participants in Round 1, 40 participants in Round 2, and 37 participants in Round 3. Experts have outlined that 25-30 participants is sufficient for Delphi studies,[36] meaning this study went above and beyond what was necessary. Finally, the fact that consensus was defined *a priori* as ≥80% agreement is noteworthy. The level of agreement in Delphi studies ranges from 51-100%,[36] meaning an 80% threshold is relatively high, and experts have stated that making this decision *a priori* reduces bias and increases the validity of the findings.[36] One of the limitations is that we only allowed those who could speak, read, and write English to participate in this study. This may have excluded some experts from non-English speaking countries. However, several members of the expert panel represented non-English speaking countries, which suggests that this may not have been the case. Another limitation is the attrition of the expert panel, as 10/58 experts (17%) did not complete Round 1, 8/48 experts (17%) did not complete Round 2, and 3/40 experts (8%) did not complete Round 3, meaning 21/58 experts (36%) did not complete all three rounds of the study. However, panel attrition is a well-known risk with Delphi studies,[36,37,76] so this was taken into account when determining the size of the expert panel,[77] and loss of 20-30% of the expert panel between rounds in Delphi studies is expected,[36] meaning the panel attrition in this study was relatively low. Finally, it must be acknowledged that 16% (n=6) of the expert panel did not agree with the long or short versions of the conceptual definition and 19% (n=7) of the expert panel did not agree with the operational definition. However, experts have pointed out that it is unlikely for an expert panel to reach 100% agreement in Delphi studies.[76]

We encourage social prescribing researchers, policymakers, and practitioners to use the definitions and framework in social prescribing research, policy, and practice. We envision several ways in which this may be done. For researchers, policymakers, and practitioners, the conceptual definitions may be used to define social prescribing in their work. For researchers conducting primary research, including qualitative, quantitative, and mixed methods studies, as well as policymakers and practitioners, the framework may be used as a guide in efforts to implement and evaluate social prescribing initiatives and policies. It may be useful to situate social prescribing initiatives and policies within the framework to ensure alignment with current understanding of this concept and to embrace a common and comparable approach. For researchers conducting evidence synthesis, such as systematic reviews, scoping reviews, and meta-analyses, the framework may be used to map the evidence base and to understand where knowledge gaps exist, and the operational definition may be used to develop inclusion and exclusion criteria. For policymakers and practitioners, the operational definition may be used to determine whether a policy or initiative meets the criteria for social prescribing. We anticipate that widespread use of the outputs of this work may help to move this field forward.

## CONCLUSION

Through a three-round Delphi study, internationally accepted conceptual and operational definitions of social prescribing were established with an international, multidisciplinary panel of experts. The CUSP conceptual framework was developed from the definitions. This foundational work offers a common thread – a shared sense of what social prescribing is. We encourage social prescribing researchers, policymakers, and practitioners to weave this common thread into social prescribing research, policy, and practice to foster common understanding of this concept.

## Data Availability

Data supporting the findings of this study are available within the manuscript and online supplemental material. Participants of this study did not agree for their individual responses to be shared publicly, so this information is not available.

## Acknowledgments

We would like to acknowledge our sincere gratitude to the expert panel.

## Competing Interests

None declared.

## Funding

The authors received no financial support for this study. The authors received financial support from the Canadian Institute for Social Prescribing for open access publishing of this article. The Canadian Institute for Social Prescribing had no involvement in the study design, data collection, data analysis, manuscript development, or the decision to submit the manuscript for publication.

## Contributors

CM conceived the study and developed the protocol in consultation with KM, IB, RA, and CG. Data collection was conducted by CM and supervised by CG. Data analysis was conducted by CM, supported by KM, and supervised by CG. CM drafted the manuscript. All authors revised the manuscript and approved the final version. CM is the guarantor of this work.

## Notes

### Competing Interest Statement

The authors have declared no competing interest.

### Author Declarations

The Health Sciences and Affiliated Teaching Hospitals Research Ethics Board at Queen's University gave ethical approval for this work.

## REFERENCES

1 Morse DF, Sandhu S, Mulligan K, et al. Global developments in social prescribing. BMJ Glob Health 2022;7:e008524. doi:10.1136/bmjgh-2022-008524

2 Global network. Global Social Prescribing Alliance. Available: https://www.gspalliance.com/global-network [Accessed 12 Oct 2022].

3 Global Social Prescribing Alliance. Social prescribing: international student movement framework, 2021. Available: https://www.gspalliance.com/student-movement-framework

4 About. Social Prescribing Network. Available: https://www.socialprescribingnetwork.com/about [Accessed 22 Apr 2022].

5 The Social Prescribing Youth Network (SPYN). Social Prescribing Network. Available: https://www.socialprescribingnetwork.com/groups-networks/special-interest-groups/spyn [Accessed 12 Oct 2022].

6 About us. National Academy for Social Prescribing. Available: https://socialprescribingacademy.org.uk/about-us/ [Accessed 22 Apr 2022].

7 Global Social Prescribing Alliance. Global Social Prescribing Alliance: international playbook, 2021. Available: https://www.gspalliance.com/gspa-playbook

8 Home. Canadian Institute for Social Prescribing. Available: https://www.socialprescribing.ca/ [Accessed 12 Oct 2022].

9 Dixon M, Polley M. Report of the annual Social Prescribing Network Conference, University of Westminster, 2016. Available: https://42b7de07-529d-4774-b3e1-225090d531bd.filesusr.com/ugd/14f499_9ba1233600eb454ab836b1c6424feed3.pdf

10 Giurca BC, Santoni CM. Celebrating the world’s first social prescribing day. Br J Gen Pract 2019;69:558. doi:10.3399/bjgp19X706325

11 World Health Organization. A toolkit on how to implement social prescribing, 2022. Available: https://www.who.int/publications/i/item/9789290619765

12 Social prescribing. World Health Organization. Available: https://openwho.org/courses/social-prescribing-WPRO [Accessed 30 May 2022].

13 Malby B, Boyle D, Wildman J, et al. The asset based health inquiry: how best to develop social prescribing, London South Bank University, 2019. Available: https://www.lsbu.ac.uk/data/assets/pdf_file/0018/251190/lsbu_asset-based_health_inquiry.pdf

14 Costa A, Sousa CJ, Seabra PRC, et al. Effectiveness of social prescribing programs in the primary health-care context: a systematic literature review. Sustainability 2021;13:2731. doi:10.3390/su13052731

15 Zurynski Y, Vedovi A, Smith K. Social prescribing: a rapid literature review to inform primary care policy in Australia, NHMRC Partnership Centre for Health System Sustainability Australian Institute of Health Innovation, Macquarie University, 2020. Available: https://chf.org.au/sites/default/files/sprapidreview_3-2-20_final.pdf

16 Rempel ES, Wilson EN, Durrant H, et al. Preparing the prescription: a review of the aim and measurement of social referral programmes. BMJ Open 2017;7:e017734. doi:10.1136/bmjopen-2017-017734

17 Islam MM. Social prescribing—an effort to apply a common knowledge: impelling forces and challenges. Front Public Health 2020;8:515469. doi:10.3389/fpubh.2020.515469

18 Bickerdike L, Booth A, Wilson PM, et al. Social prescribing: less rhetoric and more reality. A systematic review of the evidence. BMJ Open 2017;7:e013384. doi:10.1136/bmjopen-2016-013384

19 Booth A, Wilson P, Bickerdike L. Evidence to inform the commissioning of social prescribing, Centre for Reviews and Dissemination, University of York, 2015. Available: https://www.york.ac.uk/media/crd/Ev%20briefing_social_prescribing.pdf

20 Carnes D, Sohanpal R, Frostick C, et al. The impact of a social prescribing service on patients in primary care: a mixed methods evaluation. BMC Health Serv Res 2017;17:1–9. doi:10.1186/s12913-017-2778-y

21 Elston J, Gradinger F, Asthana S, et al. Does a social prescribing ‘holistic’ link-worker for older people with complex, multimorbidity improve well-being and frailty and reduce health and social care use and costs? A 12-month before-and-after evaluation. Prim Health Care Res Dev 2019;20:1–10. doi:10.1017/S1463423619000598

22 Gibbons A, Howarth M, Lythgoe A. Social prescribing in Greater Manchester, University of Salford, 2019. Available: https://usir.salford.ac.uk/id/eprint/52968/1/SocialPrescribingGMFullReport2019.pdf

23 Kimberlee R. What is social prescribing? Advances in Social Sciences Research Journal 2015;2. doi:10.14738/assrj.21.808

24 Kinsella S. Social prescribing: a review of the evidence, 2015. Available: https://www.scie-socialcareonline.org.uk/social-prescribing-a-review-of-the-evidence/r/a11G000000CTdhKIAT

25 Lejac B. A desk review of social prescribing: from origins to opportunities, Support in Mind Scotland, 2021. Available: https://www.rsecovidcommission.org.uk/a-desk-review-of-social-prescribing-from-origins-to-opportunities/

26 Moffatt S, Steer M, Lawson S, et al. Link worker social prescribing to improve health and well-being for people with long-term conditions: qualitative study of service user perceptions. BMJ Open 2017;7:e015203. doi:10.1136/bmjopen-2016-015203

27 Pescheny J, Pappas Y, Randhawa G. Facilitators and barriers of implementing and delivering social prescribing services: a systematic review. BMC Health Serv Res 2018;18. doi:10.1186/s12913-018-2893-4

28 Pescheny J, Gunn LH, Randhawa G, et al. The impact of the Luton social prescribing programme on energy expenditure: a quantitative before-and-after study. BMJ Open 2019;9:e026862. doi:10.1136/bmjopen-2018-026862

29 Pilkington K, Loef M, Polley M. Searching for real-world effectiveness of health care innovations: scoping study of social prescribing for diabetes. J Med Internet Res 2017;19:e20. doi:10.2196/jmir.6431

30 Polley M, Fleming J, Anfilogoff T, et al. Making sense of social prescribing, University of Westminster, 2017. Available: https://waystowellness.org.uk/site/assets/files/1317/making-sense-of-social-prescribing_2017_2.pdf

31 Westlake D, Elston J, Gude A, et al. Impact of COVID-19 on social prescribing across an Integrated Care System: a Researcher in Residence study. Health Social Care Comm 2022:1– 9. doi:10.1111/hsc.13802

32 Moore C, Unwin P, Evans N, et al. Social prescribing: exploring general practitioners’ and healthcare professionals’ perceptions of, and engagement with, the NHS model. Health Social Care Comm 2022. doi:10.1111/hsc.13935

33 Percival A, Newton C, Mulligan K, et al. Systematic review of social prescribing and older adults: where to from here? Fam Med Community Health 2022;10:e001829. doi:10.1136/fmch-2022-001829

34 Sandhu S, Lian T, Drake C, et al. Intervention components of link worker social prescribing programmes: a scoping review. Health Soc Care Community 2022. doi:10.1111/hsc.14056

35 Richards W. The Zen of Empirical Research. Vancouver, BC: Empirical Press 1998. Available: https://www.sfu.ca/personal/archives/richards/Zen/Pages/zentoc.html

36 Chalmers J, Armour M. The Delphi technique. In: Liamputtong P, ed. Handbook of Research Methods in Health Social Sciences. Singapore: Springer Singapore 2019:715–35. doi:10.1007/978-981-10-5251-4_99

37 Puig A, Adams C. Delphi technique. The SAGE Encyclopedia of Educational Research, Measurement, and Evaluation. 2018. doi:10.4135/9781506326139

38 Niederberger M, Spranger J. Delphi technique in health sciences: a map. Front Public Health 2020;8. doi:10.3389/fpubh.2020.00457

39 Metzelthin SF, Rostgaard T, Parsons M, et al. Development of an internationally accepted definition of reablement: a Delphi study. Ageing and Society 2020:1–16. doi:10.1017/S0144686X20000999

40 Tait AR, Geisser ME. Development of a consensus operational definition of child assent for research. BMC Med Ethics 2017;18. doi:10.1186/s12910-017-0199-4

41 Allen J, Brenner M, Hauer J, et al. Severe Neurological Impairment: a Delphi consensus-based definition. European Journal of Paediatric Neurology 2020;29:81–6. doi:10.1016/j.ejpn.2020.09.001

42 Avdagic E, Wade C, McDonald M, et al. Resilience in young children: a Delphi study to reach consensus on definitions, measurement and interventions to build resilience. Early Child Development and Care 2020;190:2066–77. doi:10.1080/03004430.2018.1556211

43 Campbell KA, Wood JN, Lindberg DM, et al. A standardized definition of near-fatal child maltreatment: results of a multidisciplinary Delphi process. Child Abuse & Neglect 2021;112:104893. doi:10.1016/j.chiabu.2020.104893

44 Beune IM, Damhuis SE, Ganzevoort W, et al. Consensus definition of fetal growth restriction in intrauterine fetal death: a Delphi procedure. Archives of Pathology & Laboratory Medicine 2021;145:428–36. doi:10.5858/arpa.2020-0027-OA

45 Ben-Chetrit E, Gattorno M, Gul A, et al. Consensus proposal for taxonomy and definition of the autoinflammatory diseases (AIDs): a Delphi study. Ann Rheum Dis 2018;77:1558–65. doi:10.1136/annrheumdis-2017-212515

46 Guseva Canu I, Marca SC, Dell’Oro F, et al. Harmonized definition of occupational burnout: a systematic review, semantic analysis, and Delphi consensus in 29 countries. Scand J Work Environ Health 2021;47:95–107. doi:10.5271/sjweh.3935

47 Dekker J, Amitami M, Berman AH, et al. Definition and characteristics of behavioral medicine, and main tasks and goals of the International Society of Behavioral Medicine—an international Delphi study. IntJ Behav Med 2021;28:268–76. doi:10.1007/s12529-020-09928-y

48 Khalil A, Beune I, Hecher K, et al. Consensus definition and essential reporting parameters of selective fetal growth restriction in twin pregnancy: a Delphi procedure. Ultrasound Obstet Gynecol 2019;53:47–54. doi:10.1002/uog.19013

49 Luquin M-R, Kulisevsky J, Martinez-Martin P, et al. Consensus on the definition of advanced Parkinson’s disease: a neurologists-based Delphi study (CEPA study). Parkinson’s Disease 2017;2017:1–8. doi:10.1155/2017/4047392

50 Monnier AA, Eisenstein BI, Hulscher ME, et al. Towards a global definition of responsible antibiotic use: results of an international multidisciplinary consensus procedure. Journal of Antimicrobial Chemotherapy 2018;73:vi3–16. doi:10.1093/jac/dky114

51 Schols AMR, Dinant G-J, Hopstaken R, et al. International definition of a point-of-care test in family practice: a modified e-Delphi procedure. Family Practice 2018;35:475–80. doi:10.1093/fampra/cmx134

52 Van den Steene H, van West D, Glazemakers I. Towards a definition of multiple and complex needs in children and youth: Delphi study in Flanders and international survey. Scandinavian Journal of Child and Adolescent Psychiatry and Psychology 2019;7:60–7. doi:10.21307/sjcapp-2019-009

53 van der Horst N, Backx F, Goedhart EA, et al. Return to play after hamstring injuries in football (soccer): a worldwide Delphi procedure regarding definition, medical criteria and decision-making. Br J Sports Med 2017;51:1583–91. doi:10.1136/bjsports-2016-097206

54 Wong HS, Curry NS, Davenport RA, et al. A Delphi study to establish consensus on a definition of major bleeding in adult trauma. Transfusion 2020;60:3028–38. doi:10.1111/trf.16055

55 Zanker J, Scott D, Reijnierse EM, et al. Establishing an operational definition of sarcopenia in Australia and New Zealand: Delphi method based consensus statement. J Nutr Health Aging 2019;23:105–10. doi:10.1007/s12603-018-1113-6

56 Adams B, Sereda M, Casey A, et al. A Delphi survey to determine a definition and description of hyperacusis by clinician consensus. International Journal of Audiology 2020:1–7. doi:10.1080/14992027.2020.1855370

57 Keller S, Salinas A, Williams D, et al. Reaching consensus on a home infusion central line-associated bloodstream infection surveillance definition via a modified Delphi approach. American Journal of Infection Control 2020;48:993–1000. doi:10.1016/j.ajic.2019.12.015

58 Lakhani BK, Giannouladis K, Leighton P, et al. Seeking a practical definition of stable glaucoma: a Delphi consensus survey of UK glaucoma consultants. Eye 2020;34:335–43. doi:10.1038/s41433-019-0540-x

59 van Helsdingen CP van, Jongen AC, Jonge WJ de, et al. Consensus on the definition of colorectal anastomotic leakage: a modified Delphi study. WJG 2020;26:3293–303. doi:10.3748/wjg.v26.i23.3293

60 Lagacé-Legendre C, Boucher V, Robert S, et al. Persistent postconcussion symptoms: an expert consensus-based definition using the Delphi method. Journal of Head Trauma Rehabilitation 2021;36:96–102. doi:10.1097/HTR.0000000000000613

61 Sudore RL, Lum HD, You JJ, et al. Defining advance care planning for adults: a consensus definition from a multidisciplinary Delphi panel. Journal of Pain and Symptom Management 2017;53:821–32. doi:10.1016/j.jpainsymman.2016.12.331

62 Bell DR, Snedden T, Biese K, et al. Consensus definition of sport specialization in youth athletes using a Delphi approach. J Athl Train 2021. doi:10.4085/1062-6050-0725.20

63 Kaur N, Pluye P. Delineating and operationalizing the definition of patient-oriented research: a modified e-Delphi study. Journal of Patient-Centered Research and Reviews 2019;6:7–16. doi:10.17294/2330-0698.1655

64 Julià-Torras J, Cuervo-Pinna MÁ, Cabezón-Gutiérrez L, et al. Definition of episodic dyspnea in cancer patients: a Delphi-based consensus among Spanish experts: the INSPIRA study. Journal of Palliative Medicine 2019;22:413–9. doi:10.1089/jpm.2018.0273

65 Feo R, Conroy T, Jangland E, et al. Towards a standardised definition for fundamental care: a modified Delphi study. J Clin Nurs 2018;27:2285–99. doi:10.1111/jocn.14247

66 D’Souza N, de Neree tot Babberich Mpm, d’Hoore A, et al. Definition of the rectum: an international, expert-based Delphi consensus. Annals of Surgery 2019;270:955–9. doi:10.1097/SLA.0000000000003251

67 Sacco S, Braschinsky M, Ducros A, et al. European headache federation consensus on the definition of resistant and refractory migraine: developed with the endorsement of the European Migraine & Headache Alliance (EMHA). J Headache Pain 2020;21. doi:10.1186/s10194-020-01130-5

68 Schaap T, Bloemenkamp K, Deneux-Tharaux C, et al. Defining definitions: a Delphi study to develop a core outcome set for conditions of severe maternal morbidity. BJOG: Int J Obstet Gy 2019;126:394–401. doi:10.1111/1471-0528.14833

69 Lawrence M, Asaba E, Duncan E, et al. Stroke secondary prevention, a non-surgical and nonpharmacological consensus definition: results of a Delphi study. BMC Res Notes 2019;12. doi:10.1186/s13104-019-4857-0

70 Casellas-Grau A, Jordán de Luna C, Maté J, et al. Developing a consensus definition of psychosocial complexity in cancer patients using Delphi methods. Pall Supp Care 2021;19:17–27. doi:10.1017/S1478951520000784

71 Katz RV, Dearing BA, Ryan JM, et al. Development of a tongue-tie case definition in newborns using a Delphi survey: the NYU–tongue-tie case definition. Oral Surgery, Oral Medicine, Oral Pathology and Oral Radiology 2020;129:21–6. doi:10.1016/j.oooo.2019.01.012

72 Smith ME, Hardman JC, Mehta N, et al. Acute otitis externa: consensus definition, diagnostic criteria and core outcome set development. PLoS One 2021;16:e0251395. doi:10.1371/journal.pone.0251395

73 Spinelli A, Anania G, Arezzo A, et al. Italian multi-society modified Delphi consensus on the definition and management of anastomotic leakage in colorectal surgery. Updates Surg 2020;72:781–92. doi:10.1007/s13304-020-00837-z

74 Yousif A, Forget A, Beauchesne M-F, et al. Development of an operational definition of treatment escalation in adults with asthma adapted to healthcare administrative databases: a Delphi study. Respiratory Medicine 2021;185:106510. doi:10.1016/j.rmed.2021.106510

75 Tack J, Boardman H, Layer P, et al. An expert consensus definition of failure of a treatment to provide adequate relief (F-PAR) for chronic constipation - an international Delphi survey. Aliment Pharmacol Ther 2017;45:434–42. doi:10.1111/apt.13874

76 Barrett D, Heale R. What are Delphi studies? Evid Based Nurs 2020;23:68–9. doi:10.1136/ebnurs-2020-103303

77 Muhl C, Mulligan K, Bayoumi I, et al. Establishing internationally accepted conceptual and operational definitions of social prescribing through expert consensus: a Delphi study protocol [Manuscript under review]. 2022.

78 Mayring P. Qualitative Content Analysis: A Step-By-Step Guide. SAGE 2021.

79 Jünger S, Payne SA, Brine J, et al. Guidance on Conducting and REporting DElphi Studies (CREDES) in palliative care: recommendations based on a methodological systematic review. Palliat Med 2017;31:684–706. doi:10.1177/0269216317690685

80 Runacres J. The problem with the definition of ‘social prescribing’: exploring the language used, and key roles within the concept, 2022. Available: http://eprints.staffs.ac.uk/7277/1/Jessica%20Runacres.%20The%20problem%20with%20th e%20definition%20of%20social%20prescribing%20Exploring%20the%20language%20used% 2C%20and%20key%20roles%20within%20the%20concept.pdf

81 Sandhu S, Alderwick H, Gottlieb LM. Financing approaches to social prescribing programs in England and the United States. Milbank Quarterly 2022;100:393–423. doi:10.1111/1468-0009.12562

82 Mulligan K. Strengthening community connections: The future of public health is at the neighbourhood scale. Toronto, ON: University of Toronto, Dalla Lana School of Public Health, 2022. Available: https://nccph.ca/projects/reports-to-accompany-the-chief-public-health-officer-of-canadas-report-2021/strengthening-community-connections-the-future-of-public-health

83 Castrucci B, Auerbach J. Meeting individual social needs falls short of addressing social determinants of health. Health Affairs, 2019. Available: https://www.healthaffairs.org/do/10.1377/hblog20190115.234942/full/ [Accessed 31 Jan 2021].

84 Green K, Zook M. When talking about social determinants, precision matters. Health Affairs, 2019. Available: https://www.healthaffairs.org/do/10.1377/hblog20191025.776011/full/ [Accessed 31 Jan 2021].

85 Kreuter MW, Thompson T, McQueen A, et al. Addressing social needs in health care settings: evidence, challenges, and opportunities for public health. Annu Rev Public Health 2021;42. doi:10.1146/annurev-publhealth-090419-102204

86 National Academies of Sciences, Engineering, and Medicine. Investing In Interventions That Address Non-Medical, Health-Related Social Needs: Proceedings of A Workshop. Washington, DC: The National Academies Press 2019.

87 Tierney S, Wong G, Mahtani KR. Current understanding and implementation of “care navigation” across England: a cross-sectional study of NHS clinical commissioning groups. Br J Gen Pract 2019;69:e675–81. doi:10.3399/bjgp19X705569

88 Husk K, Blockley K, Lovell R, et al. What approaches to social prescribing work, for whom, and in what circumstances? A realist review. Health Soc Care Community 2020;28:309–24. doi:10.1111/hsc.12839

