## Supplementary material for "Establishing Internationally Accepted Conceptual and Operational Definitions of Social Prescribing Through Expert Consensus: A Delphi Study": Online Supplemental Material 1

**Online Supplemental Material 1.** Items for the conceptual and operational definitions of social prescribing.

| **Item** | **Round 2 Results*†§ (n=40)** | **Round 2 Ratings (n=40)** | | | | |
|---|---|---|---|---|---|---|
| | | **Strongly Agree (5)** | **Agree (4)** | **Neutral (3)** | **Disagree (2)** | **Strongly Disagree (1)** |
| **Purpose** | | | | | | |
| Purpose of Social Prescribing Includes: Address Health Inequalities | ***31 (78%)*** | 17 (43%) | 14 (35%) | 5 (13%) | 2 (5%) | 2 (5%) |
| Purpose of Social Prescribing Includes: Address Non-Medical, Health-Related Social Needs | **38 (95%)** | 26 (65%) | 12 (30%) | 2 (5%) | 0 (0%) | 0 (0%) |
| Purpose of Social Prescribing Includes: Address Social Determinants of Health | **37 (93%)** | 27 (68%) | 10 (25%) | 2 (5%) | 1 (3%) | 0 (0%) |
| Purpose of Social Prescribing Includes: Improve Health and Wellbeing | **37 (93%)** | 31 (78%) | 6 (15%) | 2 (5%) | 0 (0%) | 1 (3%) |
| Purpose of Social Prescribing Includes: Reduce Health Care Demand | 22 (55%) | 8 (20%) | 14 (35%) | 11 (28%) | 6 (15%) | 1 (3%) |
| Purpose of Social Prescribing Includes: Strengthen Community Connections | **35 (88%)** | 23 (58%) | 12 (30%) | 5 (13%) | 0 (0%) | 0 (0%) |
| **People** | | | | | | |
| People Involved in Social Prescribing Include: Person Who Receives Social Prescription (Recipient) | **40 (100%)** | 33 (83%) | 7 (18%) | 0 (0%) | 0 (0%) | 0 (0%) |
| Person Who Receives Social Prescription (Recipient) is a Patient | 17 (43%) | 11 (28%) | 6 (15%) | 6 (15%) | 13 (33%) | 4 (10%) |
| Person Who Receives Social Prescription (Recipient) Has Needs | **34 (85%)** | 18 (45%) | 16 (40%) | 5 (13%) | 0 (0%) | 1 (3%) |
| Needs of Person Who Receives Social Prescription (Recipient) Cannot Be Adequately Addressed in Health Care | 29 (73%) | 14 (35%) | 15 (38%) | 6 (15%) | 5 (13%) | 0 (0%) |
| Needs of Person Who Receives Social Prescription (Recipient) Impact Health and Wellbeing | **37 (93%)** | 28 (70%) | 9 (23%) | 3 (8%) | 0 (0%) | 0 (0%) |
| Person Who Receives Social Prescription (Recipient) Receives Social Prescription for Medical Needs | 16 (40%) | 3 (8%) | 13 (33%) | 11 (28%) | 9 (23%) | 4 (10%) |
| Medical Needs Include Issues With Mental Health and Addiction, Chronic Disease, Physical Activity, etc. | 29 (73%) | 14 (35%) | 15 (38%) | 8 (20%) | 1 (3%) | 2 (5%) |
| Person Who Receives Social Prescription (Recipient) Receives Social Prescription for Non-Medical, Health-Related Social Needs | ***31 (78%)*** | 21 (53%) | 10 (25%) | 4 (10%) | 4 (10%) | 1 (3%) |
| Non-Medical, Health-Related Social Needs Include Issues With Housing, Food, Employment, Income, Social Support, etc. | **34 (85%)** | 23 (58%) | 11 (28%) | 2 (5%) | 2 (5%) | 2 (5%) |
| Person Who Receives Social Prescription (Recipient) Receives Social Prescription for (1) Medical Needs or (2) Non-Medical, Health-Related Social Needs or Both (1) and (2) | 28 (70%) | 19 (48%) | 9 (23%) | 9 (23%) | 3 (8%) | 0 (0%) |

| Item | Round 2 Results*†§ (n=40) | Round 2 Ratings (n=40) | | | | |
|---|---|---|---|---|---|---|
| | | Strongly Agree (5) | Agree (4) | Neutral (3) | Disagree (2) | Strongly Disagree (1) |
| People Involved in Social Prescribing Include: Person Who Gives Out Social Prescription (Prescriber) | **37 (93%)** | 29 (73%) | 8 (20%) | 2 (5%) | 1 (3%) | 0 (0%) |
| Person Who Gives Out Social Prescription (Prescriber) is a Health Professional | 16 (40%) | 8 (20%) | 8 (20%) | 15 (38%) | 6 (15%) | 3 (8%) |
| Person Who Gives Out Social Prescription (Prescriber) is a Health Professional or Another Authority Figure in the Community | 21 (53%) | 11 (28%) | 10 (25%) | 8 (20%) | 8 (20%) | 3 (8%) |
| People Involved in Social Prescribing Include: Person Who Gives Out Referral (Referrer) | **33 (83%)** | 24 (60%) | 9 (23%) | 4 (10%) | 3 (8%) | 0 (0%) |
| Person Who Gives Out Referral (Referrer) is a Health Professional | 23 (58%) | 9 (23%) | 14 (35%) | 8 (20%) | 6 (15%) | 3 (8%) |
| Person Who Gives Out Referral (Referrer) is Person Who Gives Out Social Prescription (Prescriber) | 25 (63%) | 11 (28%) | 14 (35%) | 6 (15%) | 6 (15%) | 3 (8%) |
| People Involved in Social Prescribing Include: Person Who Does Linking/Connecting (Linking/Connecting Agent) | **37 (93%)** | 28 (70%) | 9 (23%) | 3 (8%) | 0 (0%) | 0 (0%) |
| Person Who Does Linking/Connecting (Linking/Connecting Agent) is a Member of a Multidisciplinary Team of Professionals | 28 (70%) | 14 (35%) | 14 (35%) | 8 (20%) | 2 (5%) | 2 (5%) |
| Person Who Does Linking/Connecting (Linking/Connecting Agent) May Be Situated in Health Care or the Community | **37 (93%)** | 21 (53%) | 16 (40%) | 2 (5%) | 0 (0%) | 1 (3%) |
| Role of Person Who Does Linking/Connecting (Linking/Connecting Agent) Includes: Co-Producing Non-Medical Treatment Plan With Person Who Receives Social Prescription (Recipient)¶ | **34 (85%)** | 28 (70%) | 6 (15%) | 4 (10%) | 2 (5%) | 0 (0%) |
| Role of Person Who Does Linking/Connecting (Linking/Connecting Agent) Includes: Motivational Interviewing | **32 (80%)** | 15 (38%) | 17 (43%) | 6 (15%) | 2 (5%) | 0 (0%) |
| Role of Person Who Does Linking/Connecting (Linking/Connecting Agent) Includes: Empowering Person Who Receives Social Prescription (Recipient) to Take Greater Control of Their Own Health and Wellbeing | **36 (90%)** | 26 (65%) | 10 (25%) | 4 (10%) | 0 (0%) | 0 (0%) |
| Role of Person Who Does Linking/Connecting (Linking/Connecting Agent) Includes: Following Up With Person Who Receives Social Prescription (Recipient) | **38 (95%)** | 25 (63%) | 13 (33%) | 2 (5%) | 0 (0%) | 0 (0%) |
| Role of Person Who Does Linking/Connecting (Linking/Connecting Agent) Includes: Linking/Connecting Person Who Receives Social Prescription (Recipient) to Non-Medical Treatment¶ | **38 (95%)** | 26 (65%) | 12 (30%) | 2 (5%) | 0 (0%) | 0 (0%) |

| Item | Round 2 Results*†§ (n=40) | Round 2 Ratings (n=40) | | | | |
|---|---|---|---|---|---|---|
| | | Strongly Agree (5) | Agree (4) | Neutral (3) | Disagree (2) | Strongly Disagree (1) |
| Role of Person Who Does Linking/Connecting (Linking/Connecting Agent) Includes: Spending Time With Person Who Receives Social Prescription (Recipient) and Building Trust | **33 (83%)** | 23 (58%) | 10 (25%) | 5 (13%) | 2 (5%) | 0 (0%) |
| Role of Person Who Does Linking/Connecting (Linking/Connecting Agent) Includes: Supporting Person Who Receives Social Prescription (Recipient) to Complete Non-Medical Treatment¶ | 29 (73%) | 14 (35%) | 15 (38%) | 8 (20%) | 2 (5%) | 1 (3%) |
| Role of Person Who Does Linking/Connecting (Linking/Connecting Agent) Includes: Providing Personalized Care and Focusing on What Matters to Person Who Receives Social Prescription (Recipient) | **36 (90%)** | 26 (65%) | 10 (25%) | 3 (8%) | 1 (3%) | 0 (0%) |
| Providing Personalized Care and Focusing on What Matters to Person Who Receives Social Prescription (Recipient) Includes: Assessing the Strengths and Gifts of Person Who Receives Social Prescription (Recipient) | **32 (80%)** | 20 (50%) | 12 (30%) | 7 (18%) | 1 (3%) | 0 (0%) |
| Providing Personalized Care and Focusing on What Matters to Person Who Receives Social Prescription (Recipient) Includes: Assessing the Needs, Interests, and Goals of Person Who Receives Social Prescription (Recipient) | **38 (95%)** | 28 (70%) | 10 (25%) | 2 (5%) | 0 (0%) | 0 (0%) |
| **Properties** | | | | | | |
| Properties of Social Prescribing Include: Asset-Based Approach | 28 (70%) | 14 (35%) | 14 (35%) | 11 (28%) | 1 (3%) | 0 (0%) |
| Properties of Social Prescribing Include: Behaviour Change | **34 (85%)** | 13 (33%) | 21 (53%) | 6 (15%) | 0 (0%) | 0 (0%) |
| Properties of Social Prescribing Include: Co-Production | **36 (90%)** | 27 (68%) | 9 (23%) | 4 (10%) | 0 (0%) | 0 (0%) |
| Properties of Social Prescribing Include: Collaborative | **37 (93%)** | 27 (68%) | 10 (25%) | 3 (8%) | 0 (0%) | 0 (0%) |
| Properties of Social Prescribing Include: Collective Action | **32 (80%)** | 16 (40%) | 16 (40%) | 6 (15%) | 2 (5%) | 0 (0%) |
| Properties of Social Prescribing Include: Involves Health Care Experts and Health Care | 26 (65%) | 11 (28%) | 15 (38%) | 12 (30%) | 2 (5%) | 0 (0%) |
| Properties of Social Prescribing Include: Involves Multiple Sectors and Stakeholders | **37 (93%)** | 22 (55%) | 15 (38%) | 3 (8%) | 0 (0%) | 0 (0%) |
| Properties of Social Prescribing Include: Involves Multidisciplinary Team | 30 (75%) | 19 (48%) | 11 (28%) | 8 (20%) | 2 (5%) | 0 (0%) |
| Properties of Social Prescribing Include: Involves Social Care Experts and Social Care | 29 (73%) | 13 (33%) | 16 (40%) | 8 (20%) | 3 (8%) | 0 (0%) |
| Properties of Social Prescribing Include: Community | **38 (95%)** | 27 (68%) | 11 (28%) | 1 (3%) | 1 (3%) | 0 (0%) |
| Properties of Social Prescribing Include: Community Development | 30 (75%) | 18 (45%) | 12 (30%) | 6 (15%) | 4 (10%) | 0 (0%) |

| Item | Round 2 Results*†§ (n=40) | Round 2 Ratings (n=40) | | | | |
|---|---|---|---|---|---|---|
| | | Strongly Agree (5) | Agree (4) | Neutral (3) | Disagree (2) | Strongly Disagree (1) |
| Properties of Social Prescribing Include: Community Referral | **35 (88%)** | 24 (60%) | 11 (28%) | 3 (8%) | 1 (3%) | 1 (3%) |
| Properties of Social Prescribing Include: Community-Based | **34 (85%)** | 24 (60%) | 10 (25%) | 3 (8%) | 2 (5%) | 1 (3%) |
| Properties of Social Prescribing Include: Contextualized | 28 (70%) | 17 (43%) | 11 (28%) | 10 (25%) | 2 (5%) | 0 (0%) |
| Properties of Social Prescribing Include: Holistic Approach | **37 (93%)** | 27 (68%) | 10 (25%) | 3 (8%) | 0 (0%) | 0 (0%) |
| Properties of Social Prescribing Include: Non-Medical | ***31 (78%)*** | 18 (45%) | 13 (33%) | 6 (15%) | 3 (8%) | 0 (0%) |
| Properties of Social Prescribing Include: Alternative Option to Medicine | 18 (45%) | 9 (23%) | 9 (23%) | 12 (30%) | 7 (18%) | 3 (8%) |
| Properties of Social Prescribing Include: Non-Medical Needs | **32 (80%)** | 19 (48%) | 13 (33%) | 4 (10%) | 4 (10%) | 0 (0%) |
| Properties of Social Prescribing Include: Person-Centred Approach | **39 (98%)** | 29 (73%) | 10 (25%) | 1 (3%) | 0 (0%) | 0 (0%) |
| Properties of Social Prescribing Include: Addresses Barriers Faced by Person Who Receives Social Prescription (Recipient) | **35 (88%)** | 25 (63%) | 10 (25%) | 5 (13%) | 0 (0%) | 0 (0%) |
| Properties of Social Prescribing Include: Focuses on What Matters to Person Who Receives Social Prescription (Recipient) | **38 (95%)** | 29 (73%) | 9 (23%) | 2 (5%) | 0 (0%) | 0 (0%) |
| Properties of Social Prescribing Include: Personalized Care | **36 (90%)** | 23 (58%) | 13 (33%) | 4 (10%) | 0 (0%) | 0 (0%) |
| Properties of Social Prescribing Include: Prevention and Health Promotion | **34 (85%)** | 18 (45%) | 16 (40%) | 5 (13%) | 1 (3%) | 0 (0%) |
| Properties of Social Prescribing Include: Empowering Person Who Receives Social Prescription (Recipient) to Take Greater Control of Their Own Health and Wellbeing | **36 (90%)** | 25 (63%) | 11 (28%) | 4 (10%) | 0 (0%) | 0 (0%) |
| Properties of Social Prescribing Include: Social Capital | 27 (68%) | 16 (40%) | 11 (28%) | 12 (30%) | 1 (3%) | 0 (0%) |
| Properties of Social Prescribing Include: Social Needs | **35 (88%)** | 21 (53%) | 14 (35%) | 5 (13%) | 0 (0%) | 0 (0%) |
| Properties of Social Prescribing Include: Transformative | 27 (68%) | 16 (40%) | 11 (28%) | 10 (25%) | 3 (8%) | 0 (0%) |
| Properties of Social Prescribing Include: It is a Pathway | 26 (65%) | 17 (43%) | 9 (23%) | 11 (28%) | 3 (8%) | 0 (0%) |
| Properties of Social Prescribing Include: It is a Referral Pathway | 30 (75%) | 14 (35%) | 16 (40%) | 8 (20%) | 1 (3%) | 1 (3%) |
| Properties of Social Prescribing Include: It is an Intervention | 26 (65%) | 15 (38%) | 11 (28%) | 10 (25%) | 3 (8%) | 1 (3%) |
| **Process** | | | | | | |
| Process of Social Prescribing Includes: Activities | **37 (93%)** | 22 (55%) | 15 (38%) | 2 (5%) | 1 (3%) | 0 (0%) |
| Process of Social Prescribing Includes: Social Activities | **37 (93%)** | 21 (53%) | 16 (40%) | 2 (5%) | 1 (3%) | 0 (0%) |
| Process of Social Prescribing Includes: Community Assets and Resources | **36 (90%)** | 25 (63%) | 11 (28%) | 4 (10%) | 0 (0%) | 0 (0%) |
| Process of Social Prescribing Includes: Community Services | **34 (85%)** | 20 (50%) | 14 (35%) | 5 (13%) | 1 (3%) | 0 (0%) |
| Process of Social Prescribing Includes: Community Supports | **35 (88%)** | 25 (63%) | 10 (25%) | 5 (13%) | 0 (0%) | 0 (0%) |
| Process of Social Prescribing Includes: Non-Medical Supports and Services | **35 (88%)** | 21 (53%) | 14 (35%) | 4 (10%) | 1 (3%) | 0 (0%) |
| Process of Social Prescribing Includes: Screening | 18 (45%) | 5 (13%) | 13 (33%) | 14 (35%) | 7 (18%) | 1 (3%) |

| Item | Round 2 Results*†§ (n=40) | Round 2 Ratings (n=40) | | | | |
|---|---|---|---|---|---|---|
| | | Strongly Agree (5) | Agree (4) | Neutral (3) | Disagree (2) | Strongly Disagree (1) |
| Identifying the Non-Medical, Health-Related Social Needs of Person Who Receives Social Prescription (Recipient) | **37 (93%)** | 22 (55%) | 15 (38%) | 3 (8%) | 0 (0%) | 0 (0%) |
| Process of Social Prescribing Includes: Referral | **37 (93%)** | 26 (65%) | 11 (28%) | 3 (8%) | 0 (0%) | 0 (0%) |
| Referral is from Health Care | 15 (38%) | 8 (20%) | 7 (18%) | 21 (53%) | 3 (8%) | 1 (3%) |
| Referral is to Non-Medical Treatment¶ | ***31 (78%)*** | 18 (45%) | 13 (33%) | 7 (18%) | 2 (5%) | 0 (0%) |
| Referral is to Person Who Does Linking/Connecting (Linking/Connecting Agent) | **32 (80%)** | 16 (40%) | 16 (40%) | 5 (13%) | 3 (8%) | 0 (0%) |
| Process of Social Prescribing Includes: Social Prescription | **34 (85%)** | 22 (55%) | 12 (30%) | 6 (15%) | 0 (0%) | 0 (0%) |
| Social Prescription is Synonymous With Referral | 16 (40%) | 8 (20%) | 8 (20%) | 13 (33%) | 10 (25%) | 1 (3%) |
| Social Prescription May Be Accessed Through Self-Referral | 29 (73%) | 14 (35%) | 15 (38%) | 6 (15%) | 2 (5%) | 3 (8%) |
| Social Prescription Must Be Accessed Through Health Professional | 16 (40%) | 8 (20%) | 8 (20%) | 4 (10%) | 13 (33%) | 7 (18%) |
| Process of Social Prescribing Includes: Linking/Connecting Function | **36 (90%)** | 27 (68%) | 9 (23%) | 3 (8%) | 1 (3%) | 0 (0%) |
| Bridges the Gap Between Medical and Non-Medical Supports and Services | **34 (85%)** | 22 (55%) | 12 (30%) | 6 (15%) | 0 (0%) | 0 (0%) |
| Linking/Connecting Function Must Be Done by Person Who Does Linking/Connecting (Linking/Connecting Agent) | 18 (45%) | 8 (20%) | 10 (25%) | 21 (53%) | 0 (0%) | 1 (3%) |
| Linking/Connecting Health and Social Care | 29 (73%) | 16 (40%) | 13 (33%) | 10 (25%) | 1 (3%) | 0 (0%) |
| Linking/Connecting Person Who Receives Social Prescription (Recipient) to Non-Medical Treatment¶ | **32 (80%)** | 14 (35%) | 18 (45%) | 8 (20%) | 0 (0%) | 0 (0%) |
| Process of Social Prescribing Includes: Supporting | **37 (93%)** | 20 (50%) | 17 (43%) | 2 (5%) | 1 (3%) | 0 (0%) |
| Person Who Receives Social Prescription (Recipient) Receives Support to Complete Non-Medical Treatment¶ | 29 (73%) | 11 (28%) | 18 (45%) | 9 (23%) | 2 (5%) | 0 (0%) |
| Process of Social Prescribing Includes: Monitoring and Evaluation | **32 (80%)** | 21 (53%) | 11 (28%) | 5 (13%) | 2 (5%) | 1 (3%) |
| Feedback Loop to Person Who Gives Out Referral (Referrer) | ***31 (78%)*** | 18 (45%) | 13 (33%) | 6 (15%) | 3 (8%) | 0 (0%) |
| Following Up With Person Who Receives Social Prescription (Recipient) | **35 (88%)** | 22 (55%) | 13 (33%) | 4 (10%) | 1 (3%) | 0 (0%) |
| Measuring Outcomes | **32 (80%)** | 19 (48%) | 13 (33%) | 5 (13%) | 3 (8%) | 0 (0%) |
| Measuring Includes: Pre and Post Assessments | **32 (80%)** | 18 (45%) | 14 (35%) | 6 (15%) | 2 (5%) | 0 (0%) |
| Measuring Includes: Qualitative and Quantitative Data | **32 (80%)** | 24 (60%) | 8 (20%) | 6 (15%) | 1 (3%) | 1 (3%) |
| Outcomes Include: Impact on Health and Wellbeing | **37 (93%)** | 25 (63%) | 12 (30%) | 3 (8%) | 0 (0%) | 0 (0%) |
| Outcomes Include: Impact on Mental Health and Wellbeing | **34 (85%)** | 23 (58%) | 11 (28%) | 5 (13%) | 1 (3%) | 0 (0%) |
| Outcomes Include: Impact on Physical Health and Wellbeing | **33 (83%)** | 20 (50%) | 13 (33%) | 6 (15%) | 1 (3%) | 0 (0%) |
| Outcomes Include: Impact on Social Health and Wellbeing | **36 (90%)** | 24 (60%) | 12 (30%) | 3 (8%) | 1 (3%) | 0 (0%) |

| Item | Round 2 Results*†§ (n=40) | Round 2 Ratings (n=40) | | | | |
|---|---|---|---|---|---|---|
| | | Strongly Agree (5) | Agree (4) | Neutral (3) | Disagree (2) | Strongly Disagree (1) |
| Outcomes Include: Impact on Non-Medical, Health-Related Social Needs | **35 (88%)** | 23 (58%) | 12 (30%) | 4 (10%) | 1 (3%) | 0 (0%) |
| Outcomes Include: Impact on Life Expectancy | 14 (35%) | 8 (20%) | 6 (15%) | 16 (40%) | 6 (15%) | 4 (10%) |
| Outcomes Include: Impact on Patient Satisfaction | **33 (83%)** | 19 (48%) | 14 (35%) | 4 (10%) | 3 (8%) | 0 (0%) |
| Outcomes Include: Impact on Health Professional Satisfaction | 22 (55%) | 14 (35%) | 8 (20%) | 10 (25%) | 7 (18%) | 1 (3%) |
| Outcomes Include: Impact on Health Care Demand | ***31 (78%)*** | 13 (33%) | 18 (45%) | 6 (15%) | 3 (8%) | 0 (0%) |
| Outcomes Include: Impact on Health Care Costs | 29 (73%) | 13 (33%) | 16 (40%) | 7 (18%) | 3 (8%) | 1 (3%) |

*Round 2 Results are the sum of the participants who rated their agreement as Agree (4) or Strongly Agree (5)

†Round 2 Results that are ≥80% have been **bolded** to denote that consensus was reached

§Round 2 Results that =78% have been **bolded** and *italicized* to denote that the percentage of agreement was within 2% of the 80% threshold

¶Participants were asked to keep in mind the following statement when rating these items in the second round: 'Non-Medical Treatment' is a placeholder for the following items: Activities; Social Activities; Community Assets and Resources; Community Services; Community Supports; Non-Medical Supports and Services. You will have the opportunity to rate your agreement with these items. 'Non-Medical Treatment' will be replaced with items that achieve consensus. When rating your agreement with items that include 'Non-Medical Treatment' as an element, please remember what it represents to you.
